# Evolution of the COVID Pandemic: A Technique for Mathematical Analysis of Data

**DOI:** 10.1101/2020.05.08.20095273

**Authors:** K. Tennakone, L. Ajith DeSilva, K. J. Wimalasena, Z. J. Welchel

## Abstract

The analysis of COVID-19 statistics in different regions of the world with the intention of understanding the trend of progression of the pandemic is a task of paramount importance. Publicly available data includes cumulative number of cases, new cases each day, and the mortality. Extracting information from this data necessitates mathematical modeling. In this note a simple technique is adopted to determine the trend towards stabilized elimination of the infection, as implicated by saturation of the cumulative number of cases. Results pertaining to several representative regions of the world are presented. In several regions, evidence there to the effect that the pandemic will come to an end. The estimated saturation values of the cumulative numbers are indicated.

## 1. Introduction

A question of utmost importance to individual nations and the whole world is how the COVID-19 pandemic would progress and whether available statistics can be utilized to determine the future trend in terms of mathematical models. In this note, we present a simple technique of analyzing publicly available COVID-19 data based on the cumulative number and the new cases each day for several geopolitical regions. Analysis indicates a current general trend of cumulative COVID-19 cases approaching an equilibrium – possibly a sign that the pandemic is slowly moving towards a halt, perhaps because of the effectiveness of control measures and development of heard immunity. In few regions, slow exponential growth seems to continue.

## 2. Method and Discussion

The publicly available data on progression of COVID in different geographical regions of the world are:

1. Cumulative number of cases *N* a t the beginning of a given date (defined as at an instant of time t).
2. New cases *dN* reported at the end of each day (after a time interval *dt* = 1, therefore *dN* is effectively 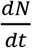). The rate of growth can also be computed from plots of *N* vs *t*.

OriginLab data analysis software is used plot curves and perform correlation analysis. The very initial growth of the epidemic in many regions has been exponential^1-2^, therefore we assume that it evolves according to the dynamical system equation,

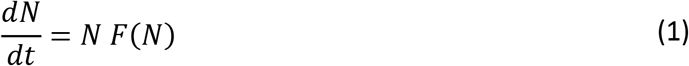

where the growth constant *F*(*N*) is a function of *N*, approximating to a constant around *N~*0. A basic assumption of the model is that the rate of increase of the number of infective (number of reported in a day) is a function of the cumulative number of cases at that instant. Implying progression of the epidemic at a given instant is governed by its history.

From (1)

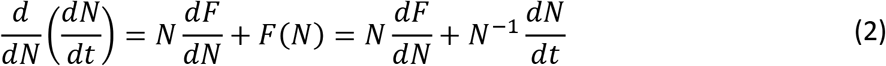

The pandemic stops brewing and reach an equilibrium when 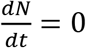 after attaining a certain cumulative number *N = N_E_*. The point *N = N_E_* is asymptotically stable^3^ at this point, provided 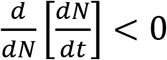. Clearly this condition satisfied if,

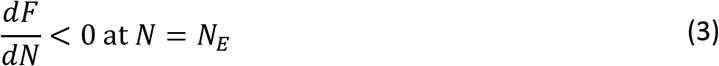

Eqn. (1) also embodies the requirement that point that point *N* = 0 corresponds to an unstable equilibrium. To illustrate these ideas, consider the logistic model, where *F*(*N*) *= α - βN*, (*α* and *β* are positive constants), with the equilibrium point 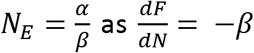, the equilibrium is stable. The cumulative number of cases in some epidemics approximately agree with the logistic model^4-7^. Our analysis shows that COVID-19 cumulative numbers deviate from the logistic variation.

The plot of *F*(*N*) vs *N* is same as the plot of 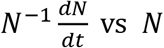, if *F*(*N*) turns out be monotonically decreasing and reaching zero, either cutting the *N* -axis or approaching *N* -axis asymptotically, the pandemic will practically reach an equilibrium. In the simple logistic model *F*(*N*) is a negative slope straight line, cutting *N* -axis at the point of equilibrium. Possible scenarios are depicted in the Fig. 1 below, which portray different modes of variation of *F*(*N*) with *N*.

**Fig. 1:**
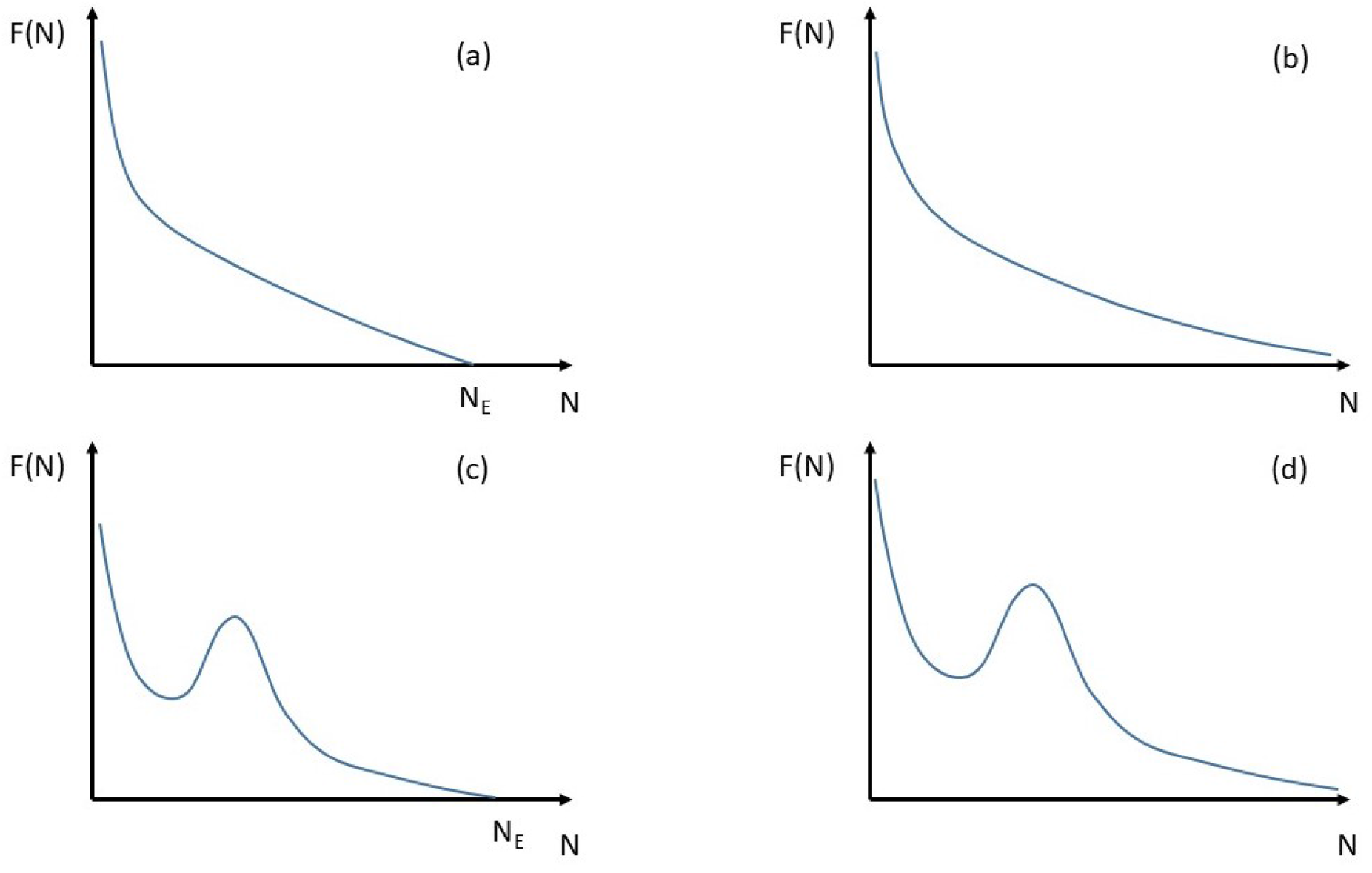
Sketch of different modes of variation *F* (*N*) with *N*.

In the figures (a) and (b) *F*(*N*) decreases continuously, and in (a) a stable equilibrium is reached at *N = N_E_*, whereas in (b) equilibrium is approached asymptomatically. In figures (c) and (d), the monotonic decrease of *N* occurs after reaching a peak. In dealing with real data, the behaviour of *F*(*N*) at points close to *N* = 0 is obviously erratic, but does interfere with the subsequent trend.

The plots of *F*(*N*) vs *N* for several representative geopolitical regions are presented below (Fig.2). All computations are based on publicly available data^8^.

**Fig. 2:**
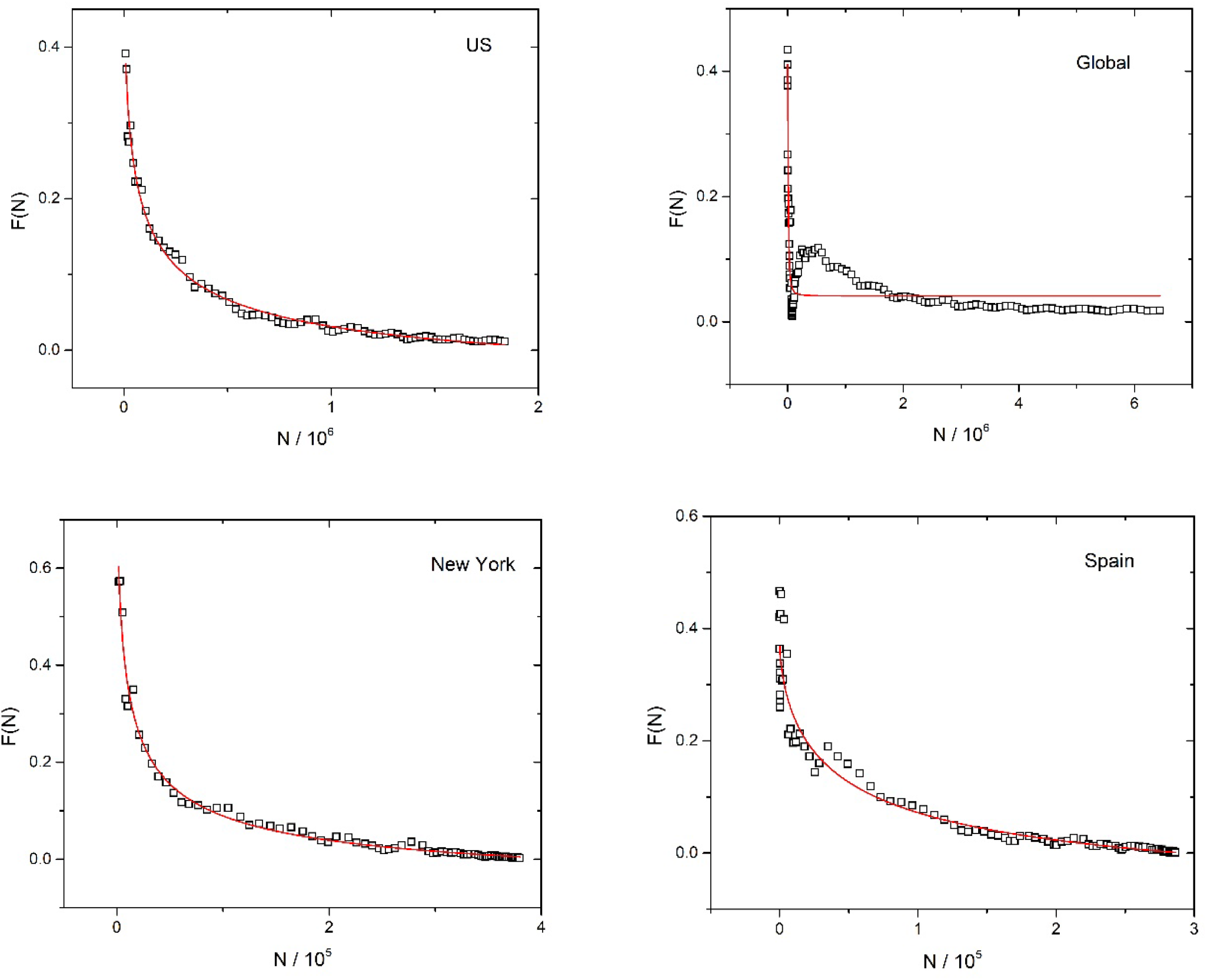
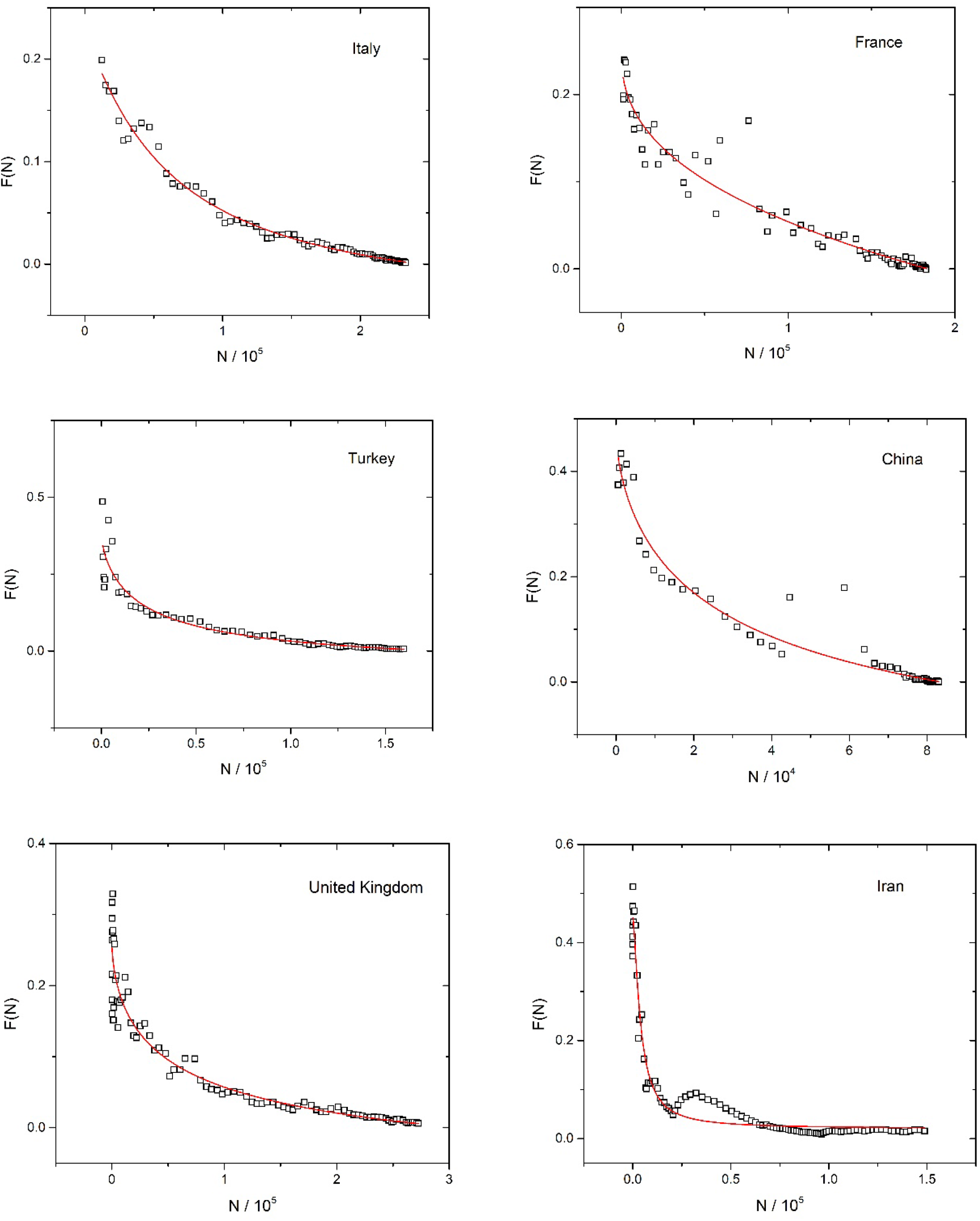
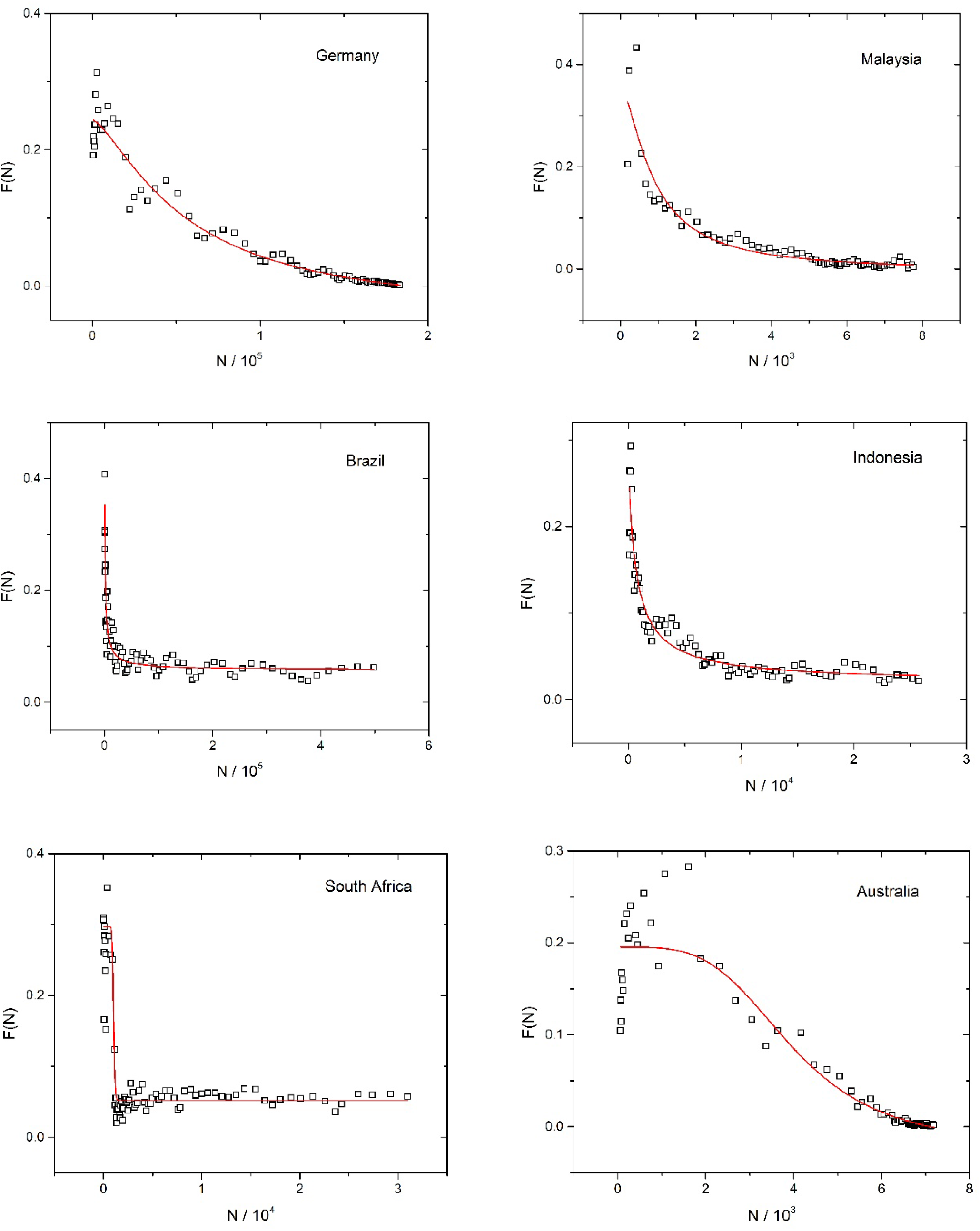
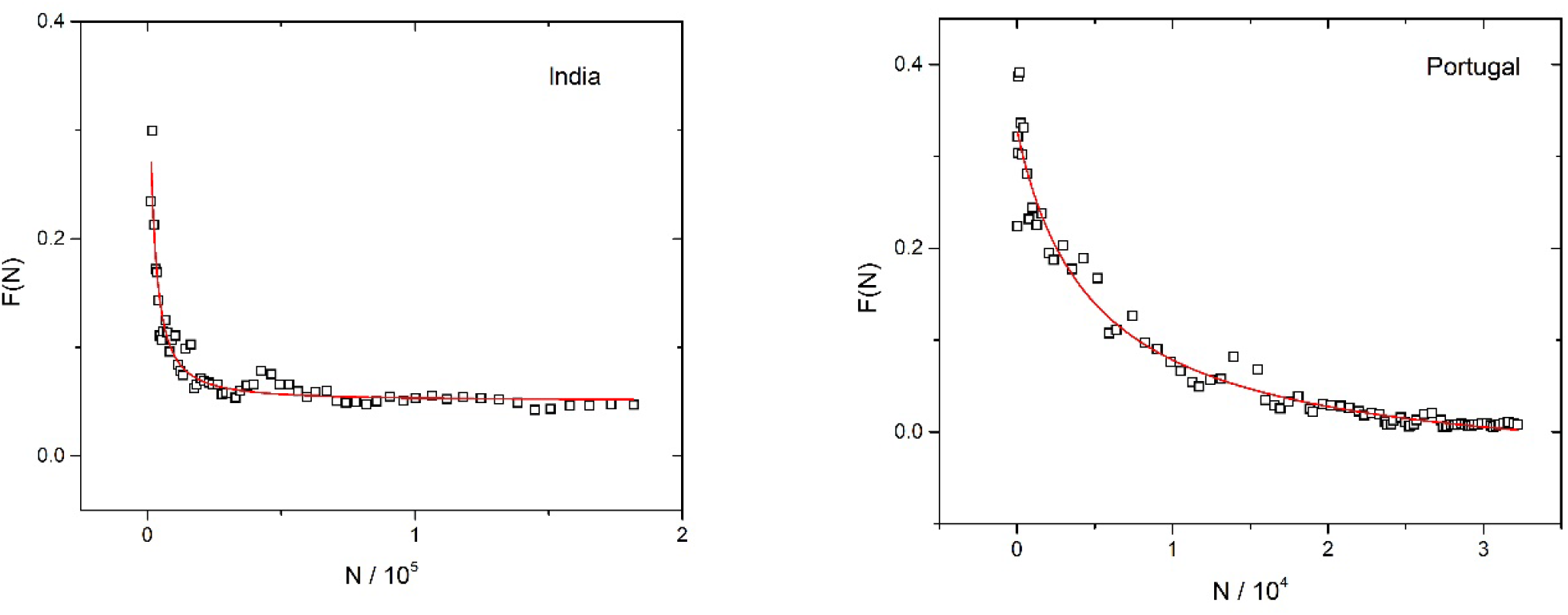
Variation of the function *F*(*N*) for different regions with number of cumulative cases *N*.

In most the above plots the later trend in variation of *F*(*N*) with *N* is monotonic decrease, indicating that 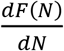 remains negative and approach *N —* a x is towards a stable equilibrium. In some regions, identification of functional form *F*(*N*) using OriginLab software, enabled determination of the point at which *F*(*N*) will cut the *N —* axis allowing determination of the saturation of the cumulative number of COVID cases. In few plots the current trend approximates to a straight line nearly parallel to *N —* axis, indicating that slow exponential growth continue.

Fig. 3 shows the plot of *F*(*N*) vs *N* for Sri Lanka. Although there is no matured COVID epidemic in Sri Lanka, the current trend is a monotonically decreasing *F*(*N*).

**Fig. 3:**
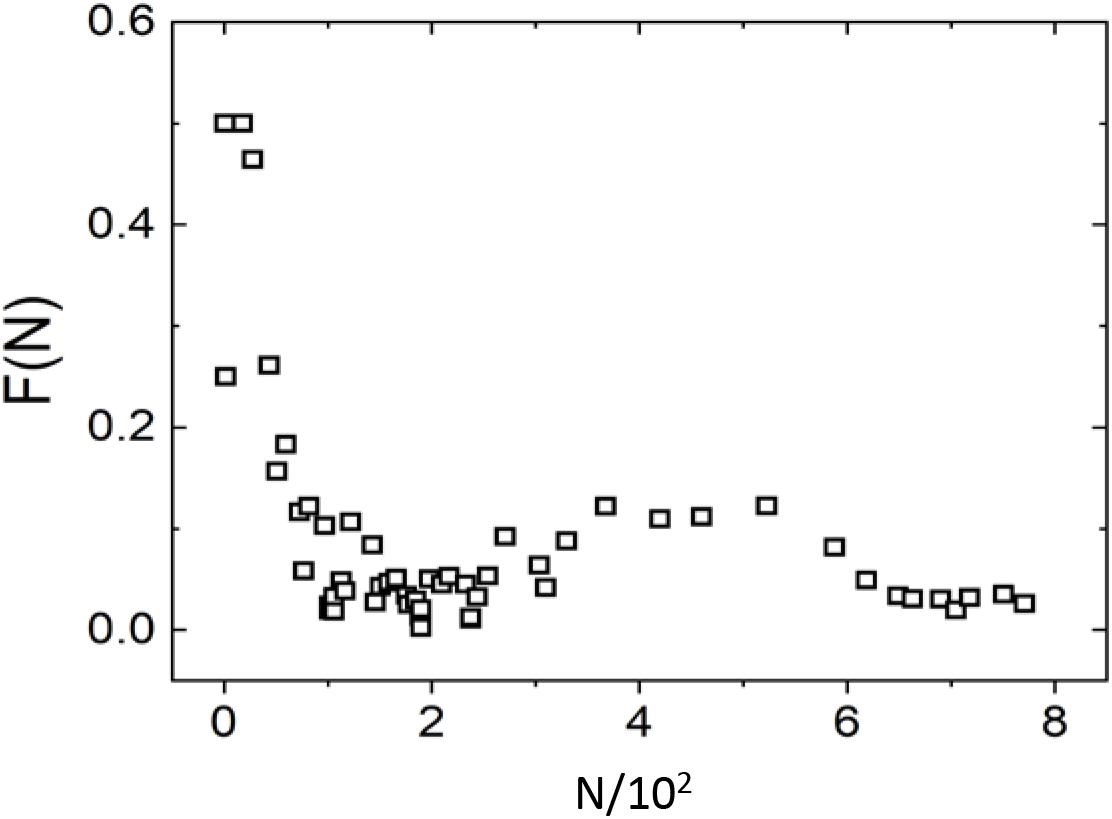
Plots of 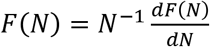 vs *N* for cumulative cases in Sri Lanka.

When curve fitting is attempted to determine the functional form of *F*(*N*), a good fit was obtained for the dose-response four parameter logistic function^9-10^,

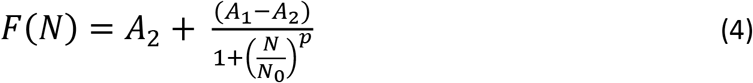

The median values of the parameters *A*_1_, *A*_2_, *N*_0_, *P* for a number of regions where the *R*^2^ confidence level in curve fitting is greater than 90% are given in the Table.1.

**Table (1):**
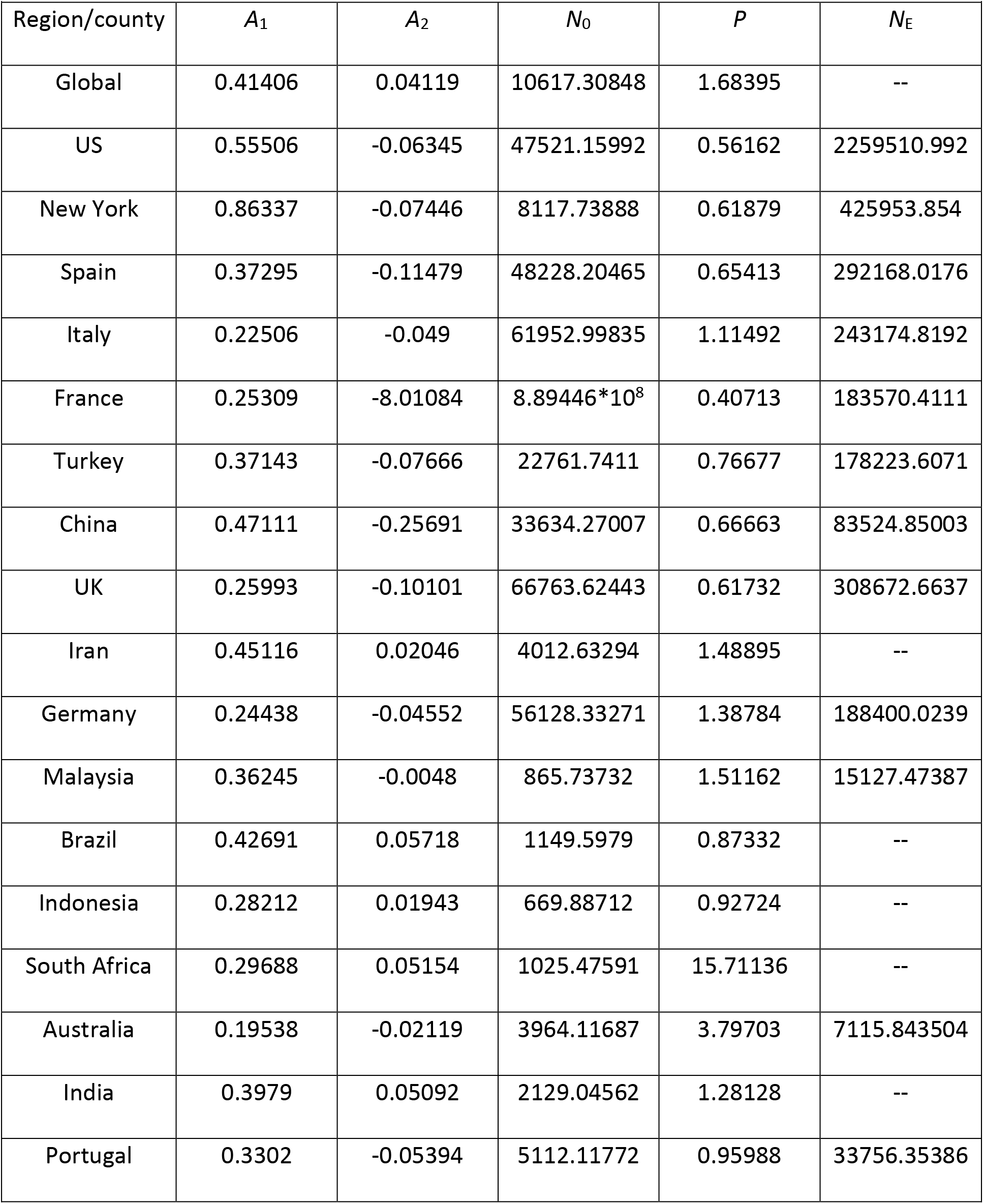
The values of the parameters *A_1_*, *A2*, *N*_0_, *P* defined in equation (4) and values of *N_E_* for different regions derived from the plots *F*(*N*) vs *N*.

In cases where *A*_2_ = (- *A*_0_) is negative, it follows from (4) that *F*(*N*) vanishes (equivalently 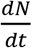) when,

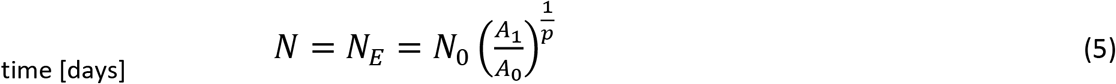

The values of *N_E_* for different regions are also indicated in Table.1. Fig.4 shows how the function *F*(*N*) for different regions vary with time. According to data analysis the value of the parameter *A_2_* is positive for Iran, Brazil, India, Indonesia and South Africa and the fitting does not suggest an approach to an equilibrium at present.

**Fig. 4:**
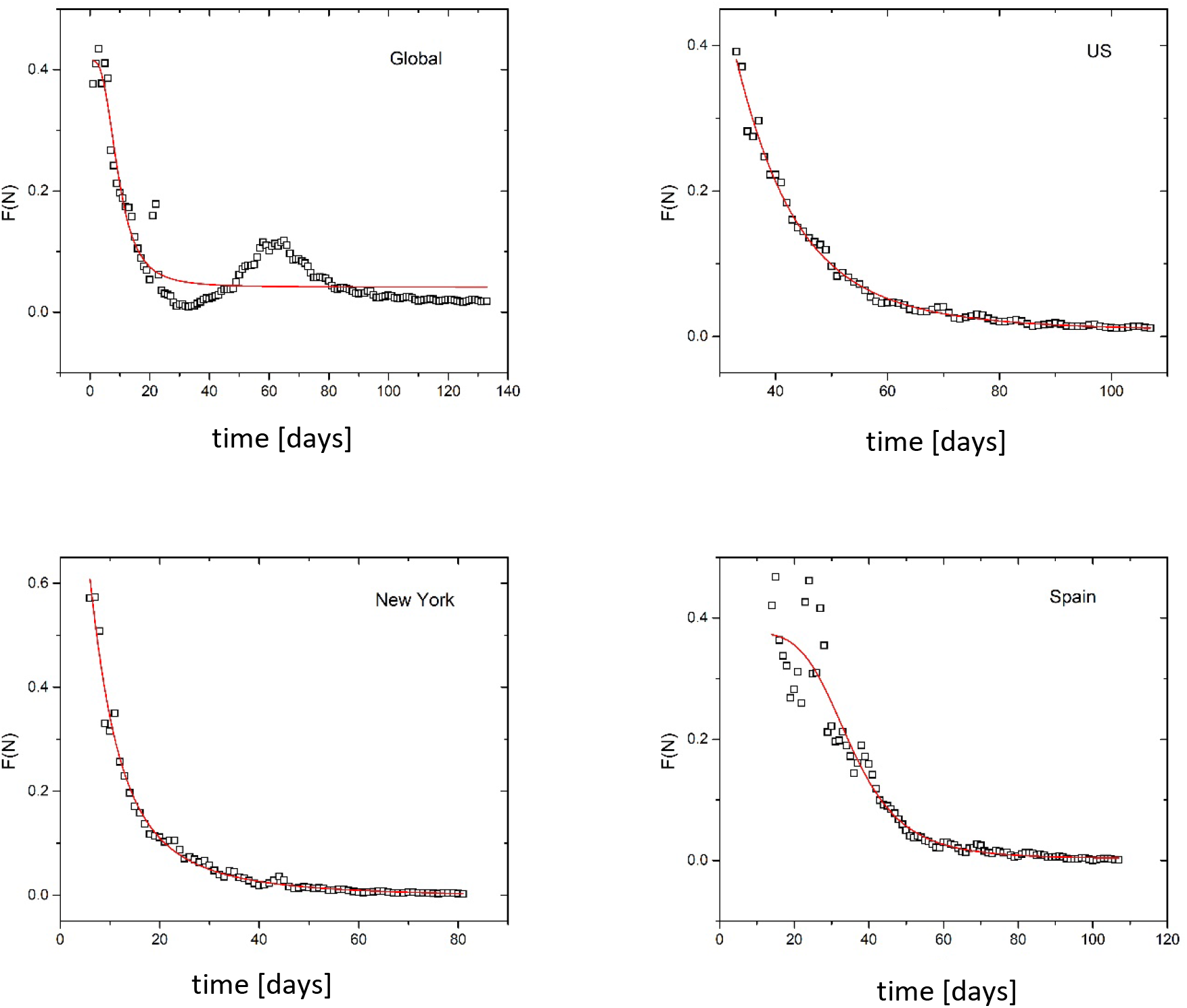
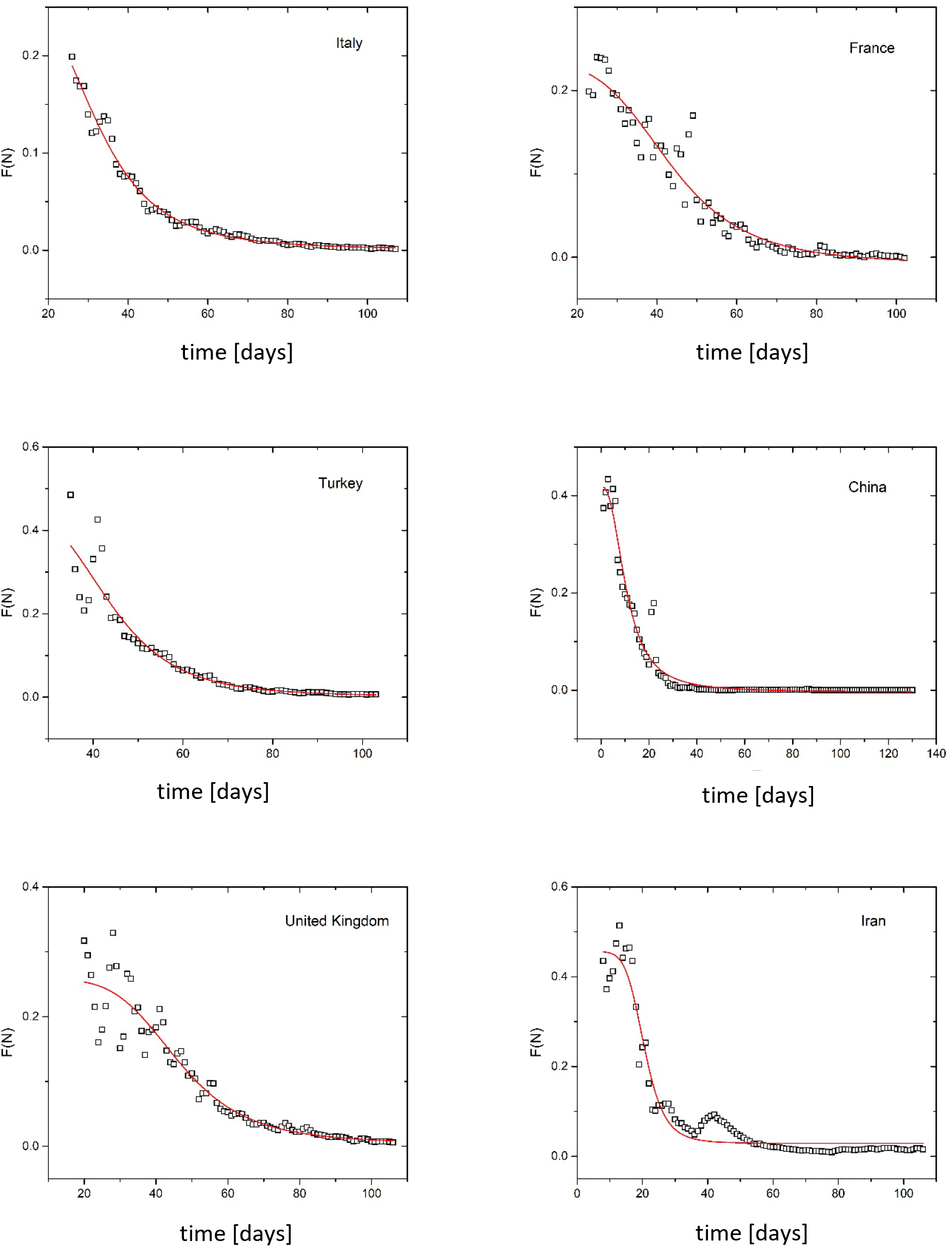
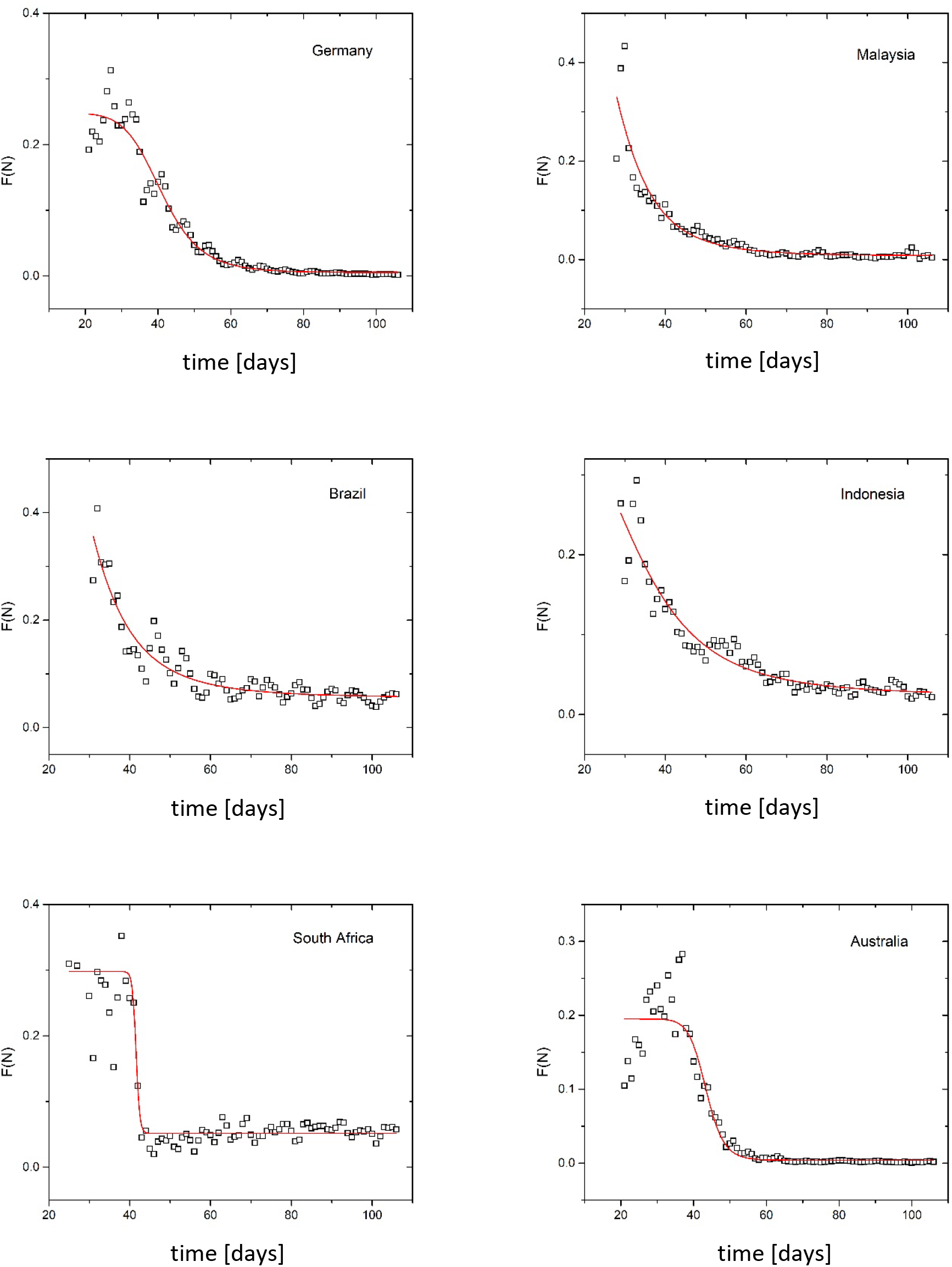
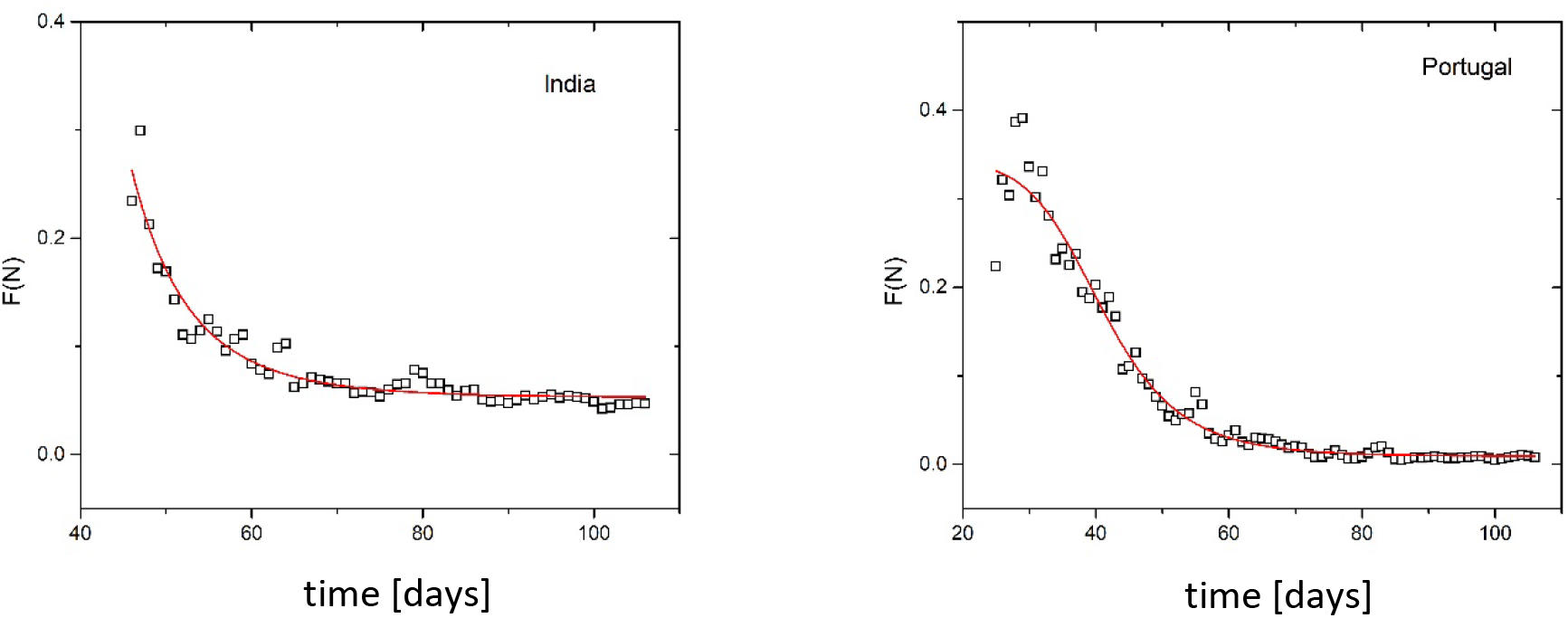
Variation of the function *F*(*N*) for different regions with time.

As seen from Fig.5, plot of ln[*F*(*N*)] vs time (*t*) fits to a straight-line after initial stages of the epidemic, so that

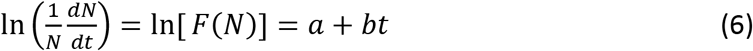

**Fig. 5:**
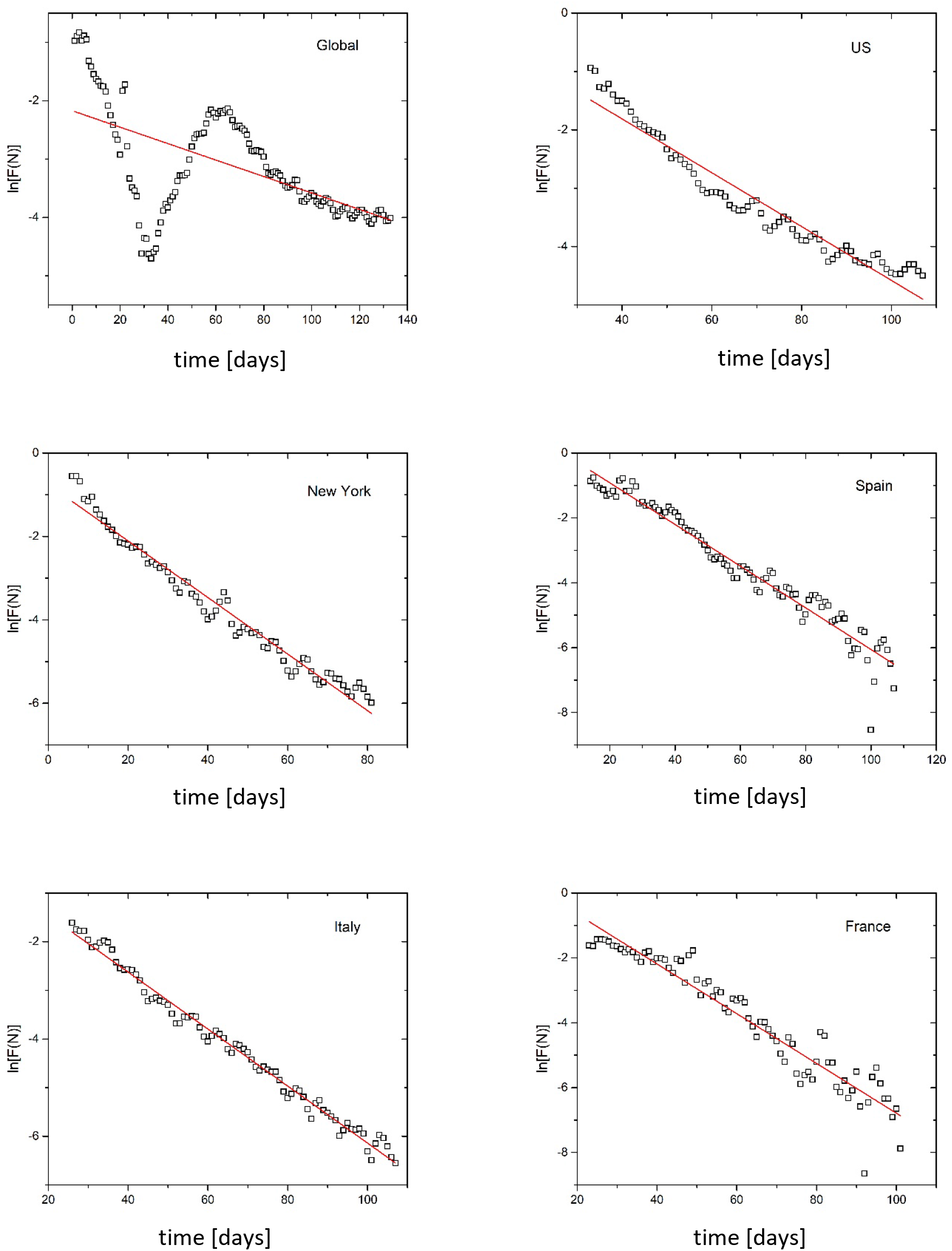
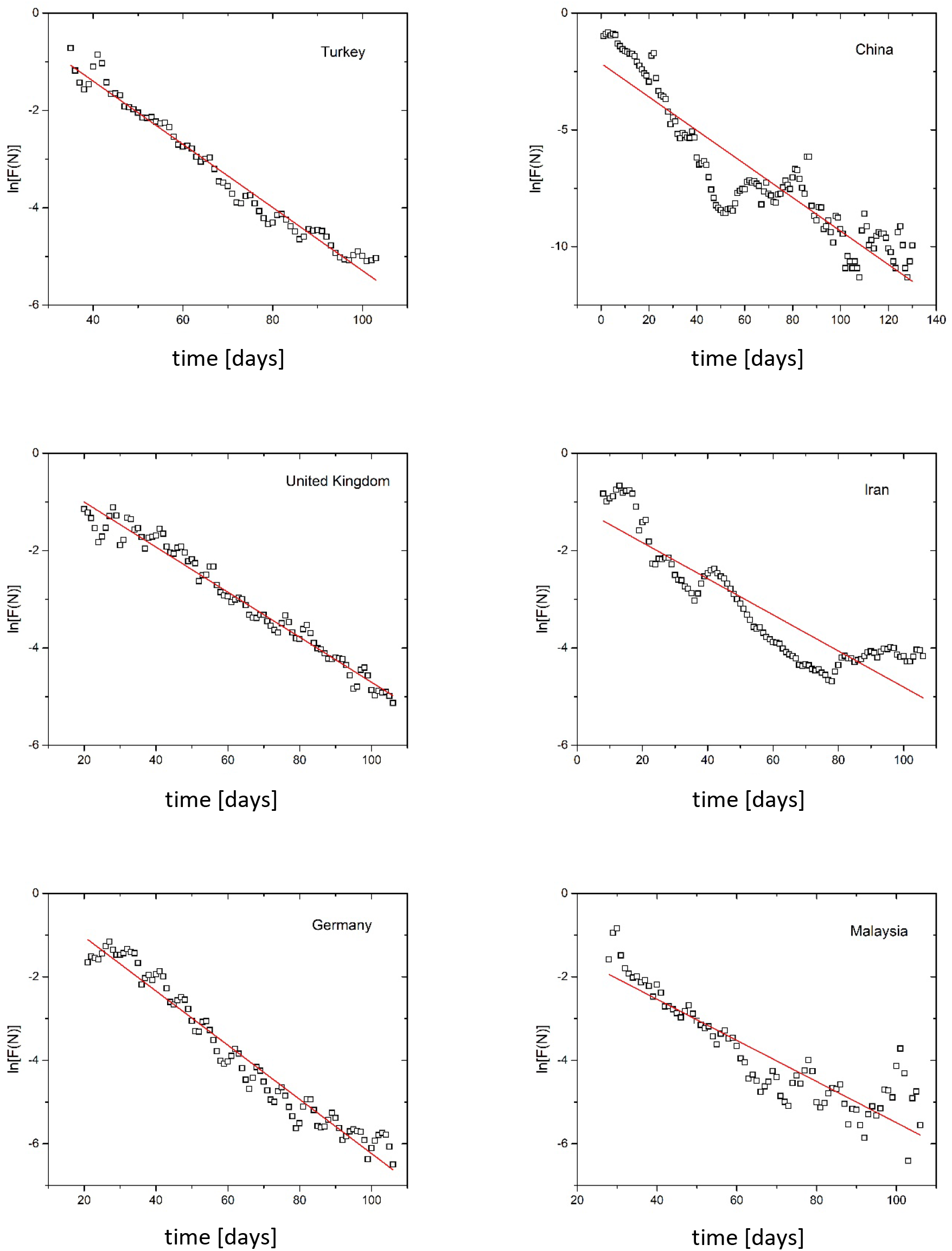
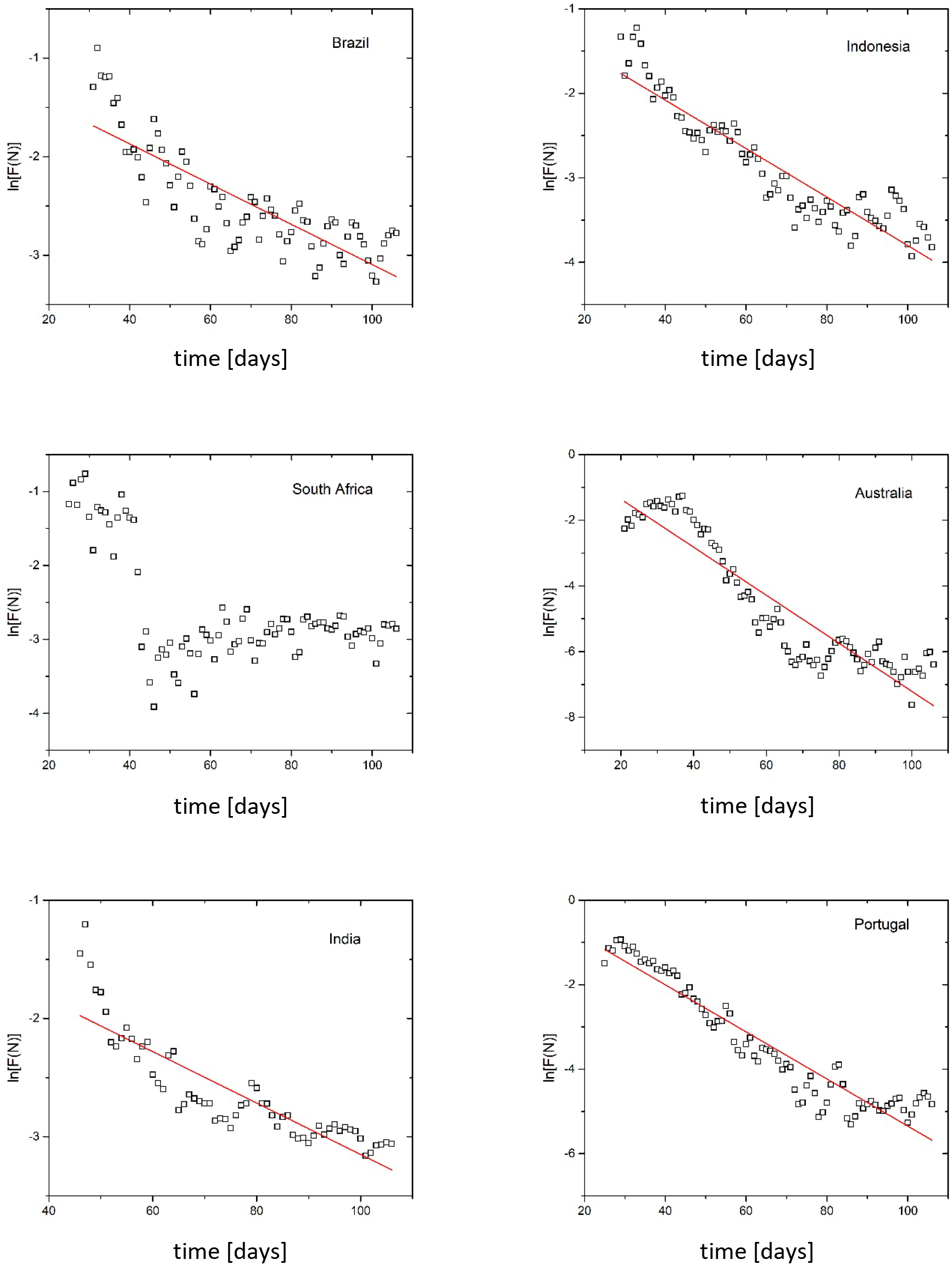
Plot of ln[*F*(*N*)] vs time.

When (6) is integrated we obtain,

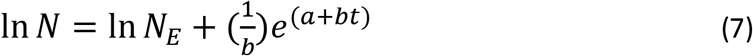

where *a* and *b* are constants.

The values of the parameters *a* and *b* are given in Table.2, parameter *b* being always negative the equilibrium *N = N_E_* is approached asymptotically and the logarithm of the cumulative number of cases decaying with a relaxation time 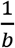 varying in the range ~ 13 — 49 days (Table 2). The value of 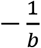 highest in Brazil and India, where the progress in achieving the equilibrium is slow and lowest in France, Australia and China. Since this is ln[N], the cumulative number of cases, approach the equilibrium very slowly. The first sketch of Figs. 2, 4 and 5 presents global data analysis. Here the value of 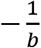 is ~ 71 days, indicating a general tendancy towards stabilization of the pandemic - the log scale cumulative numbers approaching a saturation value with relaxation time of 71 days.

**Table (2):** Values of the parameters *a* and *b* defined in eqn. (7) and value of the relaxation time *b*^-1^.

|  |  |  |  |
| --- | --- | --- | --- |
| | $a$ | $b$ | $b^{-1}(\text{days})$ |
| Global | -2.17298 | -0.01408 | 71.02273 |
| US | 0.03133 | -0.0461 | 21.69197 |
| New York | -0.75288 | -0.06779 | 14.75144 |
| Spain | 0.36705 | -0.06424 | 15.56663 |
| Italy | -0.28148 | -0.05857 | 17.07359 |
| France | 0.88323 | -0.07671 | 13.03611 |
| Turkey | 1.19755 | -0.06489 | 15.4107 |
| China | -2.15384 | -0.07175 | 13.93728 |
| UK | -0.07713 | -0.04627 | 21.61228 |
| Iran | -1.09328 | -0.03711 | 26.94691 |
| Germany | 0.25788 | -0.06492 | 15.40357 |
| Malaysia | -0.56168 | -0.04934 | 20.26753 |
| Brazil | -1.05468 | -0.0204 | 49.01961 |
| Indonesia | -0.93308 | -0.02869 | 34.85535 |
| South Africa | - | - | - |
| Australia | 0.10604 | -0.07313 | 13.67428 |
| India | -0.97645 | -0.02174 | 45.99816 |
| Portugal | 0.22618 | -0.05572 | 17.94688 |

## Conclusion

The analysis suggests that fitting COVID-19 cumulative cases *N* to an equation of the form 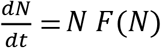 gives insight into the progression pandemic in different regions. As 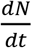 is essentially the number of cases reported each day, the function *F*(*N*) can be easily plotted. Since 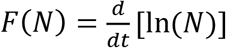, this quantity can be readily ascertained from logarithmic plots of the cumulative number. The variation of *F*(*N*) with *N* depicts the progression towards stabilization of the cumulative number. If *F*(*N*) approaches *N* —axis with negative slope, indication is that the equilibrium point *N_E_* within reach. This tendency is visible in majority of the curves presented - most probably a signature of success of the containment measures.

Forecasting the progress of the pandemic is possible by fitting data to determine the functional form of *F*(*N*) and best fit was obtained for the four parameter dose-response logistic function given by eqn.(4). Analysis enabled determination of saturation value of the cumulative number for most regions.

More information about the evolution of the pandemic could follow on updating of the plots. A limitation of the model is assumption of the equation (1) where *F*(*N*) not an explicit function of *t*. Dynamical systems interpretations, apply only under the above assumption. Relaxation of control measures, mutations of the virus or influences of the climate may alter the observed trends *F*(*N*) forecasted here.

## Data Availability

Public available data:
All Data Retrieved from JHU Data Repository:(Last access 06/03/2020)

https://github.com/CSSEGISandData/COVID-19/blob/master/csse_covid_19_data/csse_covid_19_time_series/time_series_covid19_confirmed_global.csv

## Disclaimer

Authors are not responsible for authenticity of data used, any errors that had in occurred in calculations or interpretations and making decisions based on this study.

## Acknowledgement

Authors wish to thank Prof. D.Tantrigoda, University of Sri Jayewardhanapura, Sri Lanka for valuable discussions, constructive comments and reading the manuscript.

